# Facemask shortage and the coronavirus disease (COVID-19) outbreak: Reflection on public health measures

**DOI:** 10.1101/2020.02.11.20020735

**Authors:** Huailiang Wu, Jian Huang, Casper J. P. Zhang, Zonglin He, Wai-Kit Ming

**Affiliations:** Department of Public Health and Preventive Medicine, School of Medicine, Jinan University, Guangzhou, China; International School, Jinan University, Guangzhou, China; MRC Centre for Environment and Health, Department of Epidemiology and Biostatistics, School of Public Health, St Mary’s Campus, Imperial College London, Norfolk Place, London W2 1PG, United Kingdom; School of Public Health, LKS Faculty of Medicine, The University of Hong Kong, Hong Kong, China; HSBC Business School, Peking University, Shenzhen, China

**Keywords:** coronavirus, COVID-19, SARS-CoV-2, facemask shortage, public health measures, health policy

## Abstract

**Background:** A novel coronavirus disease (COVID-19) outbreak due to SARS-CoV-2 infection occurred in China in late-December 2019. Facemask wearing is considered as one of the most cost-effective and important measures to prevent the transmission of SARS-CoV-2, but it became a social concern due to the recent global facemask shortage. China is the major facemask producer in the world, contributing to 50% of global production. However, even full productivity (20 million facemasks per day) does not seem to meet the need of a population of 1.4 billion in China.

**Methods:** Policy review using government websites and shortage analysis using mathematical modelling based on data obtained from the National Health Commission (NHC), the Ministry of Industry and Information Technology (MIIT), the Center for Disease Control and Prevention (CDC) of the People’s Republic of China, and Wuhan Bureau of Statistics.

**Findings:** Supplies of facemasks in the whole of China would have been sufficient for both healthcare workers and the general population if the COVID-19 outbreak only occurred in Wuhan city or Hubei province. However, if the outbreak occurred in the whole of China, facemask supplies in China could last for 5 days if under the existing public health measures and a shortage of 853 million facemasks is expected by 30 Apr 2020. Assuming a gradually decreased import volume, we estimated that dramatic increase in productivity (42.7 times of the usual level) is needed to mitigate the facemask crisis by the end of April.

**Interpretation:** In light of the COVID-19 outbreak in China, a shortage of facemasks and other medical resources can considerably compromise the efficacy of public health measures. Effective public health measures should also consider the adequacy and affordability of medical resources. Global collaboration should be strengthened to prevent the development of a global pandemic from a regional epidemic via easing the medical resources crisis in the affected countries.

**Research in context:** *Evidence before this study:* We searched PubMed and Web of Science for articles in English, between 1 Jan 1980, and 1 Jan 2020, using the search terms 1) (infection OR infectious disease* OR outbreaks) AND (modelling); and 2) (mask* OR facemask* OR medical resource*) AND (infection OR infectious disease* OR outbreaks). Most relevant studies identified were performed to predict diseases spread and to determine the original infection source of previous epidemics like SARS and H7N9. However, few studies focused on the medical resources crisis during the outbreaks.

*Added value of this study:* To the best of our knowledge, this is the first study to investigate the facemask shortage during the novel coronavirus pneumonia (COVID-19) outbreak in China. We have summarized in detail the management strategies implemented by the Chinese governments during the outbreaks. By considering three scenarios for the outbreak development, we simulated the facemasks availability from late-December 2019 to late-April 2020 and estimated the duration of sufficient facemask supplies. Our findings showed that if the COVID-19 outbreak occurred only in Wuhan city or Hubei province, facemask shortage would not appear with the existing public health measures. However, if the outbreak occurred in the whole of China, a shortage of facemask could be substantial assuming no alternative public health measures.

*Implications of all the available evidence:* Our findings provide insight into the public health measures to confront medical resources crisis during infectious disease outbreaks. Effective public health measures should consider the adequacy and affordability of existing medical resources. Governments across the world should revisit their emergency plans for controlling infectious disease outbreaks by taking into account the supply of and demand for the medical resource. Global collaboration should be strengthened to prevent the development of a global pandemic from a regional epidemic via easing the medical resources crisis in the affected countries.

## Introduction

An increasing number of novel coronavirus disease (COVID-19) cases were initially identified in Wuhan, Hubei province, China in December 2019. The novel coronavirus (SARS-CoV-2) was mainly transmitted via respiratory droplets and can be transmitted between humans ^1-3^. Common symptoms of COVID-19 include fever, cough, dyspnoea, and myalgia or fatigue, while less common symptoms include sputum production, headache, haemoptysis, and diarrhoea ^4^. In mid-February 2020, the reported incidence of COVID-19 cases exceeded 60,000 in China, of which more than 70% was in Wuhan city and more than 80% was in Hubei province ^5,6^. Globally, Thailand, Japan, South Korea, Singapore, Malaysia, France, Canada, Australia, Germany, the United Kingdom, the United States and 13 other countries had reported COVID-19 cases ^7^. Most of the confirmed cases are travellers from or ever been to Wuhan or other Chinese cities; however, locally transmitted cases outside of China have also been reported ^7^.

The World Health Organization’s (WHO) guidance on prevention and control of the COVID-19 outbreak recommends hand, and respiratory hygiene and the use of appropriate personal protective equipment for healthcare workers in practice and patients with suspected SARS-CoV-2 infection should be offered a medical mask ^8^. Regarding the respiratory hygiene measures, facemask wearing is considered as one of the most cost-effective and important measures to prevent the transmission of SARS-CoV-2, but it became a social concern due to the recent global facemask shortage ^9-12^.

China is the major facemask producer in the world, contributing to 50% of the global production ^9^. At the usual time, China can produce 20 million facemasks per day while the productivity during the Chinese New Year holiday was lower (12 million facemasks per day) ^13^. However, even full productivity does not seem to meet the need of a population of 1·4 billion in China. Therefore, to control the COVID-19 epidemic, the Chinese government imported more than 730 million facemasks between 24 Jan 2020 and 11 Feb 2020 ^13,14^ and extended the Chinese New Year holiday to allow for home quarantine and to reduce the need for facemasks and other medical resources. A number of factories also resumed partial productivity during the holiday by paying extra to their workers ^15^.

In this study, we simulated the facemask availability during the COVID-19 outbreak using a mathematical model based on the actual development of the outbreak, public health measures introduced by the Chinese government, and national statistics on facemask production and import. We aim to investigate the situation of the facemask shortage during the COVID-19 outbreak in China and reflect on the existing public health measures. This analysis could provide insight into the development of emergency plans regarding the adequacy and affordability of medical resources for future infectious disease outbreaks.

## Method

A cluster of COVID-19 cases was reported by the Wuhan Municipal Health Commission in late Dec 2019 ^1^, and the peak of the epidemic is predicted to be between mid-to-late-February based on data from Wuhan while the epidemic is predicted to fade out within two months after the peak ^16,17^. The number of new cases is expected to decline after the epidemic peak, but the viral transmission is still possible and the need for facemask will not decrease immediately. Therefore, our analysis covered the period from 31 Dec 2019 to 30 Apr 2020 (121 days in total). To simulate the facemask availability in China, we used a mathematical model based on data and assumptions on the production, import, and need. We considered three scenarios in which the COVID-19 outbreak occurred in 1) Wuhan city (the epicenter); 2) Hubei province (the province/state where the epicenter is located); and 3) the whole of China (the entire country).

### Mathematical model

We simulated the daily facemasks availability using equation (1),

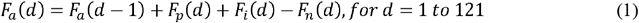

where *F*_*a*_*(d)* is the facemasks availability, as defined as the total number of facemasks on the market and in storage at the end of day *d. F*_*a*_*(0)* (i.e., *d=1*) is the baseline facemask availability at the beginning of our prediction period. *F*_*p*_*(d)* is the number of facemasks produced in China on day *d. F*_*i*_*(d)* is the number of facemasks imported to China on day *d. F*_*n*_*(d)* is the need for facemasks on day *d*.

We estimated the daily need for facemasks using equations (2) to (4),

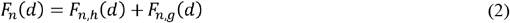

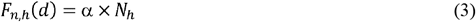

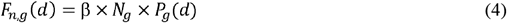

where *F*_*n,h*_*(d)* and *F*_*n,g*_*(d)* are the need for facemasks among healthcare workers and the general population on day *d* in the disease epidemic region, respectively. *α* and *β* are the daily need for facemasks of one healthcare worker and one person in the general population in the disease epidemic region, respectively. *N*_*h*_ and *N*_*g*_ are the numbers of healthcare workers and the general population in the disease epidemic region, respectively. *P*_g_(*d*) is the percentage of the general population in the disease epidemic region need facemasks on day *d*.

### Data source and model assumptions

Public health measures introduced by the Chinese government during the COVID-19 outbreak were summarized based on the official announcements and documents from the National Health Commission (NHC), the Ministry of Industry and Information Technology (MIIT), and the Center for Disease Control and Prevention (CDC) of the People’s Republic of China, and Municipal Health Commission.

The parameters included in the mathematical model were estimated based on the data released by the NHC, MIIT, and CDC of the People’s Republic of China or recent investigation on the outbreak.Model input and assumptions in our model are summarized in Table 1. Specifically, the population and the numbers of healthcare workers in Wuhan city, Hubei province, and China were obtained from the Wuhan Bureau of Statistics and China Health Statistics Yearbook 2019 ^18-20^. MIIT estimated that the daily facemask production in China is about 20 million at usual time ^13^. Facemask storage was estimated to be seven times the daily consumption in hospitals ^9^. We assume that 70% of the facemask storage in the whole of China would be supplied to the hospital system; thus, the baseline facemasks availability (*F*_*a*_*(0)*) was estimated to be 246,006,500, which is about 12 times of the usual daily facemask productivity in China. In our model, we assumed all facemasks on the market and in storage are available for consumption. In other words, we did not take into account factors that may limit the supply on the market, such as logistics. We assumed that the daily facemask productivity (*F*_*p*_*(d)*) changed during our prediction period due to the Chinese New Year holiday (24 Jan 2020 to 9 Feb 2020), the incentive to resume production, and new product lines. Specifically, we considered three scenarios for the changes of daily facemask productivity, i.e., 1) 100% productivity (20 million per day) before the Chinese New Year holiday, 40% to 60% productivity during the holiday, and 94% to 100% productivity after the holiday; 2) 100% productivity before the Chinese New Year holiday, 40% to 60% productivity during the holiday, and the productivity would increase from 94% after the holiday to 200% on 3 Mar 2020; 3) 100% productivity before the Chinese New Year holiday, 40% to 60% productivity during the holiday, and the productivity would increase from 94% to 400% on 3 Mar 2020 (See Table 1 for detailed description of each scenario). According to the MIIT and General Administration of Customs, the daily number of facemasks imported to China were 6,000,000 between 24 Jan 2020 and 29 Jan 2020, 20,000,000 on 30 Jan 2020^13^, 56,166,667 between 31 Jan 2020 and 11 Feb 2020^14^. Given a global facemask shortage has appeared in mid-February, we assumed a gradually decreasing daily import of facemask between 12 Feb 2020 and 23 Mar 2020 from 50% to 10% of the import volume in early-February and maintained a 10% of the import volume in early-February from 23 Mar 2020 to 30 Apr 2020. Based on the clinical experience of chief physicians in China, we assumed that each healthcare worker would use five facemasks each day (*α*) at the usual time. And we assumed that each person in the general population would use one facemask each day (*β*). We assumed the percentage of the general population wearing facemasks (*P*_*g*_*(d)*) to be close to 0% before the release of the national technical protocol for SARS-CoV-2 (the first version) by the NHC (i.e., 0<*d*≤16) ^2^. The *P*_*g*_*(d)* was assumed to be 20% after the NHC recommended facemasks wearing in the national technical protocol (the first version) (16<*d*≤23), and 60% after the release of the national technical protocol for SARS-CoV-2 (the second version) and massive media cover (23<*d*≤25) ^21^. The Chinese government suggested home quarantine for suspected cases and reducing gathering during the Chinese New Year holiday; thus, we assumed the *P*_*g*_*(d)* to be 20% from 24 Jan 2020 to 9 Feb 2020 (25<*d*≤41). Given that most employees would return to work on 10 Feb 2020 while students would return to schools on 2 Mar 2020^22^, we assumed the *P*_*g*_*(d)* to be 40% from 10 Feb 2020 to 1 Mar 2020 (41<*d*≤61) and 60% from 1 Mar 2020 to 30 Apr 2020 (61<*d*≤121).

**Table 1.** Parameters for estimating the facemask availability during the novel coronavirus disease (COVID-19) outbreak in China.

| Parameter | Definition | Data source or assumptions | Model input |
| --- | --- | --- | --- |
| $N_{p,c}$ | Population of mainland China | Data from National Health Commission of the People's Republic of China | 1,395,380,000 |
| $N_{p,h}$ | Population of Hubei province | Data from National Health Commission of the People's Republic of China | 59,170,000 |
| $N_{p,w}$ | Population of Wuhan city | Data from Wuhan Bureau of Statistics | 11,080,000 |
| $N_{h,c}$ | Number of healthcare workers in mainland China | Data from National Health Commission of the People's Republic of China | 12,300,325 |
| $N_{h,h}$ | Number of healthcare workers in Hubei province | Data from National Health Commission of the People's Republic of China | 521,930 |
| $N_{h,w}$ | Number of healthcare workers in Wuhan city | Data from National Health Commission of the People's Republic of China and Wuhan Bureau of Statistics | 145,209 |
| $P_g(d)$ | Percentage of population using facemasks on day $d$ | Between 31 Dec 2019 and 15 Jan 2020: 0%<br>Between 16 Jan 2020 and 22 Jan 2020: 20%<br>Between 23 Jan 2020 and 24 Jan 2020: 60%<br>Between 25 Jan 2020 and 9 Feb 2020: 20%<br>Between 10 Feb 2020 and 1 Mar 2020: 40%<br>Between 2 Mar 2020 and 30 Apr 2020: 60% | 0% ($0 < d \leq 16$)<br>20% ($16 < d \leq 23$)<br>60% ($23 < d \leq 25$)<br>20% ($25 < d \leq 41$)<br>40% ($41 < d \leq 61$)<br>60% ($61 < d \leq 121$) |
| $\alpha$ | Number of facemasks consumption of each healthcare worker each day | Assumed that each healthcare workers would use five facemasks each day | 5 |
| $\beta$ | Number of facemasks consumption of each person in the general population each day | Assumed that each person in the general population would use one facemask each day | 1 |
| $F_p(d)$ | Number of facemasks produced in China on day $d$ | Estimation based on information from Ministry of Industry and Information Technology of the People's Republic of China<br>**Scenario 1:**<br>Between 31 Dec 2019 and 23 Jan 2020: 20,000,000<br>Between 24 Jan 2020 and 1 Feb 2020: 8,000,000 | **Scenario 1:**<br>20,000,000 ($0 < d \leq 24$)<br>8,000,000 ($24 < d \leq 33$) |
|  |  | Between 2 Feb 2020 and 10 Feb 2020: 12,000,000<br>Between 11 Feb 2020 and 17 Feb 2020: 18,800,000<br>Between 18 Feb 2020 and 30 Apr 2020: 20,000,000<br>**Scenario 2:**<br>Between 31 Dec 2019 and 23 Jan 2020: 20,000,000<br>Between 24 Jan 2020 and 1 Feb 2020: 8,000,000<br>Between 2 Feb 2020 and 10 Feb 2020: 12,000,000<br>Between 11 Feb 2020 and 17 Feb 2020: 18,800,000<br>Between 18 Feb 2020 and 24 Feb 2020: 20,000,000<br>Between 25 Feb 2020 and 2 Mar 2020: 30,000,000<br>Between 3 Mar 2020 and 30 Apr 2020: 40,000,000<br>**Scenario 3:**<br>Between 31 Dec 2019 and 23 Jan 2020: 20,000,000<br>Between 24 Jan 2020 and 1 Feb 2020: 8,000,000<br>Between 2 Feb 2020 and 10 Feb 2020: 12,000,000<br>Between 11 Feb 2020 and 17 Feb 2020: 18,800,000<br>Between 18 Feb 2020 and 24 Feb 2020: 20,000,000<br>Between 25 Feb 2020 and 2 Mar 2020: 40,000,000<br>Between 3 Mar 2020 and 30 Apr 2020: 80,000,000 | 12,000,000 ($33 < d \leq 41$)<br>18,800,000 ($41 < d \leq 48$)<br>20,000,000 ($48 < d \leq 121$)<br>**Scenario 2:**<br>20,000,000 ($0 < d \leq 24$)<br>8,000,000 ($24 < d \leq 33$)<br>12,000,000 ($33 < d \leq 41$)<br>18,800,000 ($41 < d \leq 48$)<br>20,000,000 ($48 < d \leq 55$)<br>30,000,000 ($55 < d \leq 62$)<br>40,000,000 ($62 < d \leq 121$)<br>**Scenario 3:**<br>20,000,000 ($0 < d \leq 24$)<br>8,000,000 ($24 < d \leq 33$)<br>12,000,000 ($33 < d \leq 41$)<br>18,800,000 ($41 < d \leq 48$)<br>20,000,000 ($48 < d \leq 55$)<br>40,000,000 ($4 < d \leq 62$)<br>80,000,000 ($62 < d \leq 121$) |
| $F_i(d)$ | Number of facemasks imported to China on day $d$ | Estimation based on information from Ministry of Industry and Information Technology of the People's Republic of China.<br>Between 24 Jan 2020 and 29 Jan 2020: 6,000,000<br>30 Jan 2020: 20,000,000<br>Between 31 Jan 2020 and 11 Feb 2020: 56,166,667 | 6,000,000 ($24 \leq d \leq 29$)<br>20,000,000 ($d=30$)<br>56,166,667 ($30 < d \leq 42$) |
|  |  | Between 12 Feb 2020 and 21 Feb 2020: 28,083,334<br>Between 22 Feb 2020 and 2 Mar 2020: 22,466,667<br>Between 3 Mar 2020 and 12 Mar 2020: 16,850,000<br>Between 13 Mar 2020 and 22 Apr 2020: 11,233,33<br>Between 23 Mar 2020 and 30 Apr 2020: 5,616,667 | 28,083,334 (42< d ≤ 52)<br>22,466,667 (52< d ≤ 62)<br>16,850,000 (62< d ≤ 72)<br>11,233,333 (72< d ≤ 82)<br>5,616,667 (82< d ≤ 121) |
| $F_a(0)$ | Number of facemasks available in the whole of China on 30 Dec 2010 | Facemask storage was estimated to be seven times the daily consumption in hospitals<sup>9</sup>. We assume that 70% of the facemask storage in the whole of China would be supplied to the hospital system; thus, $F_a(0) = 7 \times \alpha \times N_{hc} / 70\% = 172,204,550 / 70\% = 246,006,500$ | 246,006,500 |

**Table 2.** Summary of recommendations for confronting medical resource crisis during infectious disease outbreaks

| Recommendations |  |
| --- | --- |
| **National level** | *Before infectious disease outbreak* |
|  | 1. Revisit emergency plans for controlling infectious disease outbreaks to include strategies to avoid medical resources shortage during the outbreaks. |
|  | *During infectious disease outbreak* |
|  | 1. Closely monitor the supply of and demand for medical resources; |
|  | 2. Timely response to the increasing demand for medical resources by increasing domestic productivity and global imports; |
|  | 3. Maintain the availability and affordability of medical resources; |
|  | 4. Reduce demand for medical resources by implementing alternative public health measures such as transport shutdown, school closure, and arrangement for effective home quarantine. |
| **International level** | *Before infectious disease outbreak* |
|  | 1. Strengthen global collaboration for confronting medical resource crisis; |
|  | 2. Establish an information sharing platform monitoring global productivity and storage of medical resource. |
|  | *During infectious disease outbreak* |
|  | 1. Evaluate the severity of the outbreak and availability of medical resource in the affected countries; |
|  | 2. Evaluate the availability of medical resources in the global collaborative network and assist the epidemic areas to maintain a sufficient supply; |
|  | 3. Give recommendation for global supply, demand and distribution if an international medical resource crisis is anticipated; |
|  | 4. Release travel advice and guidance of self-protection accounting for potential medical resource crisis. |

## Results

Figure 1 shows the cumulative numbers of confirmed cases and deaths of the COVID-19 and the public health measures introduced by the Chinese government from 31 Dec 2019 to 15 Feb 2020, e.g., identification of pathogen, activation of different levels of emergency response, announcement of protocols and guidelines for healthcare workers and the general population to follow, legislation facemask wearing in public, and lockdown of about 13 cities in Hubei province. From municipal level, provincial level, to national level, the Chinese government had implemented various strategies to control and prevent the spread of COVID-19.

**Figure 1.**
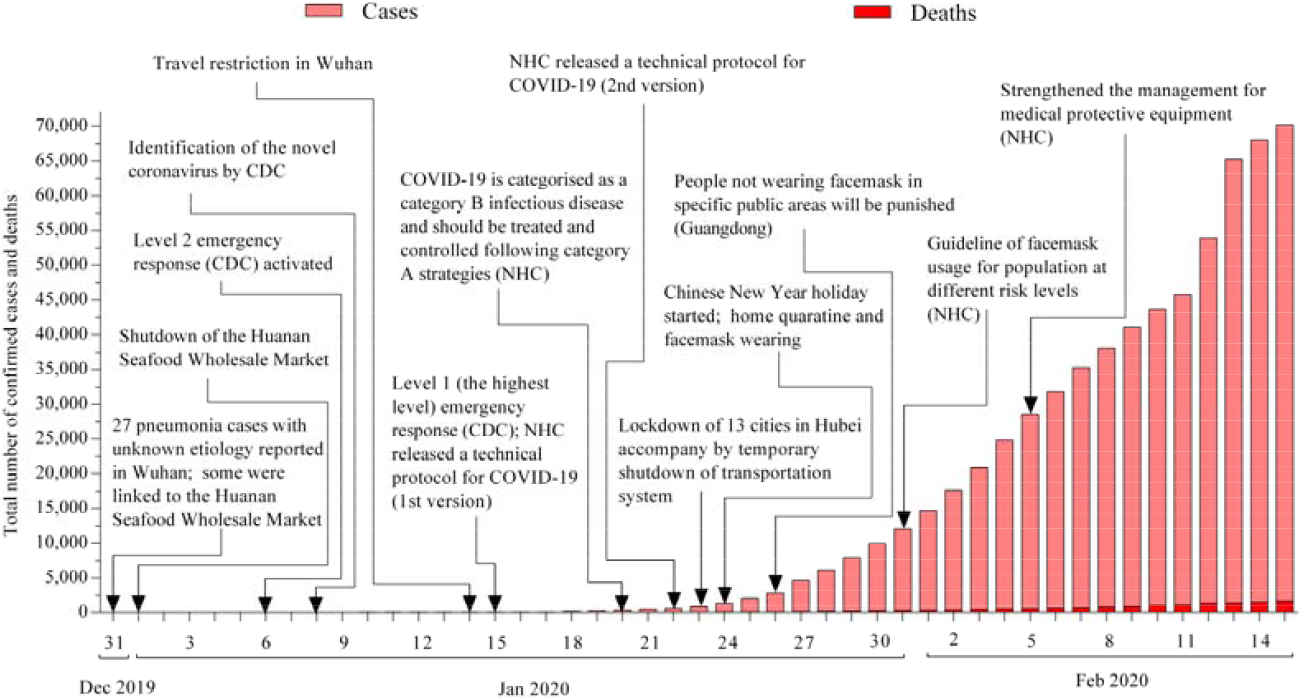
Timeline of public health measures introduced by the Chinese government during the novel coronavirus disease (COVID-19) outbreak. (Publicly available data from official websites of the National Health Commission (NHC), the Ministry of Industry and Information Technology (MIIT), and the Center for Disease Control and Prevention (CDC) of the People’s Republic of China)

Figure 2 shows our simulation of the facemask availability during the COVID-19 outbreak under different scenarios of outbreak scale and facemask productivity. Specifically, Figure 2a shows the facemask availability during the outbreak if the productivity only resumed to the usual level after the Chinese New Year holiday. In this scenario, if the COVID-19 outbreak only occurred in Wuhan city or Hubei province (Scenarios a1 and a2), facemasks supplies in the whole of China could meet the need in these regions. However, if the COVID-19 outbreak occurred in the whole of China (Scenario a3), facemasks availability could only last for about 5 days (from 31 Dec 2019 to 5 Jan 2020). On 30 April, there would be a shortage of approximate 853 million in the whole of China. Figures 2b and 2c show the facemask availability during the outbreak if facemask productivity can gradually increase to 200% and 400% of the usual level, respectively. As expected, if the COVID-19 outbreak only occurred in Wuhan city or Hubei province (Scenarios b1, b2, c1, and c2), facemasks supplies in the whole of China could meet the need in these regions. However, even with a productivity of 400% of the usual level, facemask shortage could not be resolved by the end of April if the COVID-19 outbreak occurred in the whole of China. We estimated that facemask productivity equals to 42.7 times the usual level is needed to mitigate a shortage by the end of April.

**Figure 2.**
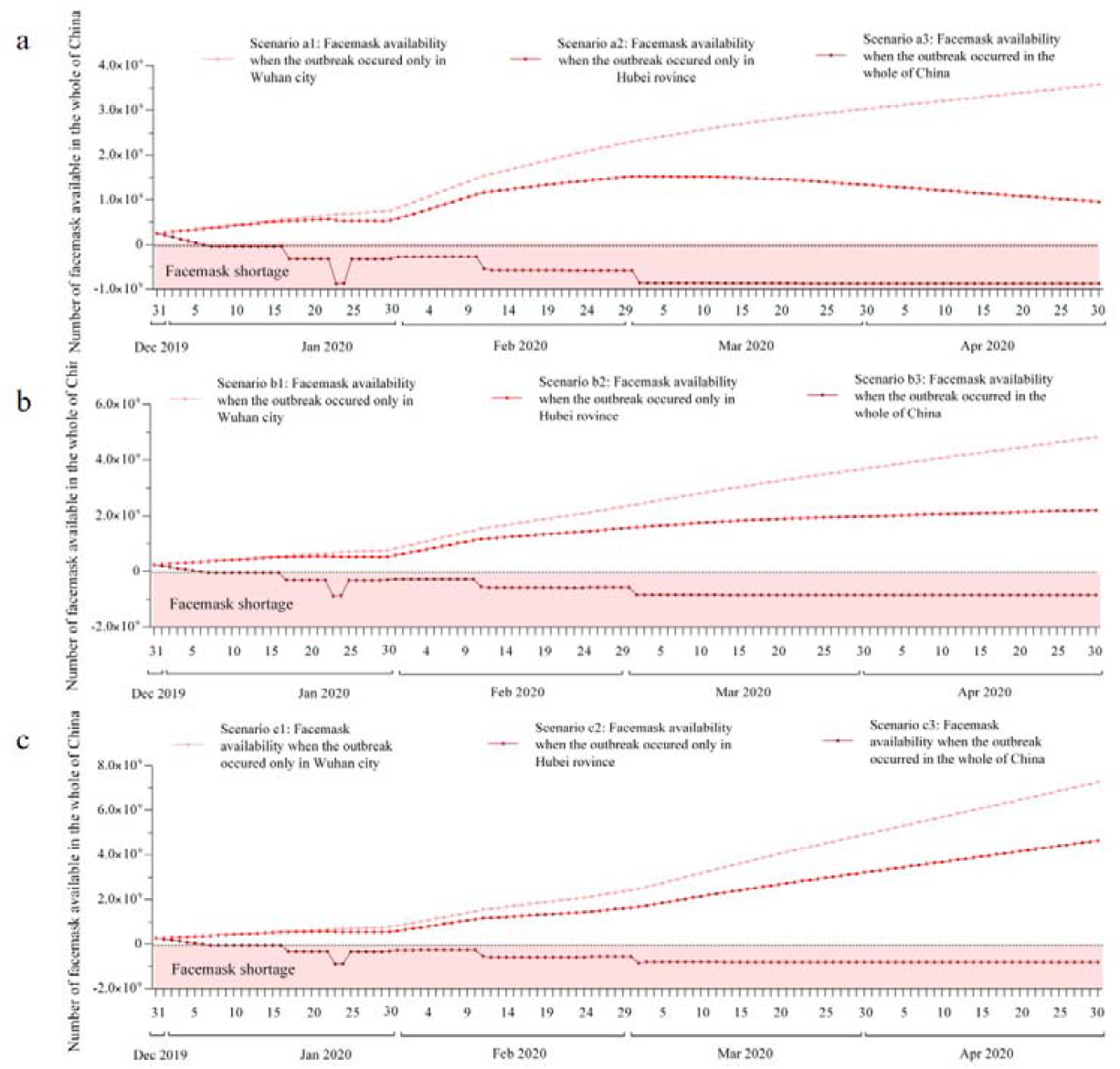
Facemask availability during the novel coronavirus disease (COVID-19) outbreak in China (a: assuming domestic facemask productivity resumed to 100% of the usual level; b: assuming domestic facemask productivity gradually increased to 200% of the usual level; c: assuming domestic facemask productivity gradually increased to 400% of the usual level)

## Discussion

In this study, we summarized the public health measures introduced by the Chinese government during the COVID-19 outbreak and simulated the facemask availability in three scenarios based on the scale of the outbreak. Our results suggested that if the COVID-19 outbreak only occurred in Wuhan (about million population) or Hubei province (about 59.2 million population), a facemask shortage would not happen in China. We also simulated the facemask availability in three scenarios regarding different facemask productivity during the outbreak. Our analysis showed that if the outbreak occurs in the whole of China, a shortage of facemask would appear in less than a week and could be substantial under the existing public health measures. Only a dramatic increase in productivity (42.7 times the usual level) could mitigate the facemask shortage in time.

During the COVID-19 outbreak, the Chinese government introduced various measures, such as lockdown of cities, shutdown of the transportation system, and legislation on mandatory facemasks wearing in public ^23^. However, the implementation of these responses was not always timely, and the virus eventually spread to the entire country. As soon as the general population began to realise the severity of the outbreak, facemask consumption surged in only a few days, partly due to panic buying. Although domestic productivity has resumed after the Chinese New Year holiday and global imports have increased, facemask crisis was not resolved. Under these circumstances, many avoided leaving their apartments and tried to extend the lifespan of each facemask by all means ^24,25^, school was closed, online learning resources became widely available, and public facilities were mostly not opened. In this way, the demand for facemasks in the general population is expected to decline and more resources could be saved for healthcare workers ^26^.

Although our analysis was based on the COVID-19 outbreak in China, our findings could provide insight into the public health measures in other areas of the world, considering the outbreak occurs in city with a relatively high population mobility (e.g., London, New York, and Tokyo), and the virus could spread to the province/state where this city is located and even the entire country quickly. Therefore, governments across the world should revisit their emergency plan for controlling infectious disease outbreaks in the local context. Timely public health measures should be taken to control the outbreak within the city or the province/state where the city is located. Meanwhile, the supply of and demand for facemasks and other medical resources should be considered when planning for public health measures, so as to maintain the availability and affordability of medical resources. Besides, timely and effective communication with the public is essential to mitigate panic buying and anxiety in the population ^27,28^. Furthermore, during a medical resource crisis, health disparity could be widened between specific population groups. Individuals of lower socioeconomic status are more likely to find themselves in a dilemma between the need to work in high-risk locations and the lack of protective equipment. In addition, market forces can drive the price up, preventing them from purchasing an adequate amount of protective equipment.

In order to prevent the development of a global pandemic from a regional epidemic, a global collaboration to ease the medical resources crisis in the affected countries during an infectious disease outbreak should be established. With the shared information, the collaboration could promptly evaluate the severity of the outbreak and the availability of medical resources. Travel advice and guidance of self-protection should also account for the potential medical resouce crisis in the epidemic areas. Timely actions, such as increasing global productivity and distributing global storage, could be taken with a joint effort to minimize the risk of shortage and control the outbreak.

To the best of our knowledge, this is the first study to investigate the facemask availability during the COVID-19 outbreak in China. We have summarized in detail the public health measures introduced by the Chinese governments and considered three scenarios for the outbreak development. Nevertheless, there are some limitations in this study. First, our estimation relied on the assumptions of facemask productivity, import, storage, and need. Relevant information was limited at the time when the analyses were performed (during the outbreak). Second, we did not take into account the logistics cost that could restrict the supplies on the markets. Thus, our analysis is likely to underestimate the severity of the facemask shortage experiencing by the healthcare workers and the general population. Third, we didn’t consider the types of facemasks that should be used by individuals at different risk levels ^29^. The situation of facemasks shortage might be more severe when considering the demand for different types of facemasks from healthcare workers (most required KN95/N95 respirators and medical protective masks) and the general population ^29^. Fourth, our prediction ended in late-April considering the epidemic was predicted to fade out at about the same time ^30^. However, if the transmissibility of the epidemic could be reduced, the peak and the end would be delayed ^31^, the daily incidence and the demand for facemasks and other medical supplies would also decrease. Nevertheless, the anxiety in the population may result in constant demand for facemasks even when the epidemic is under controlled.

## Conclusion and policy implication

In light of the COVID-19 outbreak in China, a shortage of facemasks and other medical resources can considerably compromise the efficacy of public health measures. Effective public health measures should also consider the adequacy and affordability of medical resources. Global collaboration should be strengthened to prevent the development of a global pandemic from a regional epidemic via easing the medical resources crisis in the affected countries.

## Data Availability

Data available on request from the authors

## Data sharing statement

All data used in this study are publicly available. Data sources are described in the method section.

## Contributor statement

| Author | Contribution |
| --- | --- |
| Dr Huailiang Wu | 1. Analysed and interpreted the data;<br>2. Visualizing the results;<br>3. Drafted the manuscript;<br>4. Revised the manuscript. |
| Dr Jian Huang | 1. Interpreted the data;<br>2. Reviewed the manuscript for intellectual content<br>3. Revised the manuscript. |
| Dr Casper J. P. Zhang | 1. Reviewed the manuscript for intellectual content<br>2. Revised the manuscript |
| Dr Zonglin He | 1. Project administration<br>2. Data curation |
| Dr Wai-Kit Ming | 1. Designed and conceptualized study;<br>2. Interpreted the data;<br>3. Reviewed the manuscript for intellectual content<br>4. Responsible for the overall content as a guarantor |

## Reference

1. Nishiura H, Jung SM, Linton NM, et al. The Extent of Transmission of Novel Coronavirus in Wuhan, China, 2020. J Clin Med 2020; 9(2).

2. Li Q, Guan X, Wu P, et al. Early Transmission Dynamics in Wuhan, China, of Novel Coronavirus-Infected Pneumonia. N Engl J Med 2020.

3. Cui J, Li F, Shi ZL. Origin and evolution of pathogenic coronaviruses. Nat Rev Microbiol 2019; 17(3): 181–92.

4. Huang C, Wang Y, Li X, et al. Clinical features of patients infected with 2019 novel coronavirus in Wuhan, China. Lancet 2020.

5. Wuhan Municipal Health Commission. Wuhan Municipal Health Commission briefing on the pneumonia epidemic situation, 13 February 2020 (in Chinese). 2020. http://wjw.hubei.gov.cn/fbjd/dtyw/202002/t20200214_2027187.shtml (accessed 14 Feb 2020.

6. National Health Commission of the People ‘s Republic of China. Updates on pneumonia of new coronavirus infections as of February 13, 2020 (in Chinese). 2020. http://www.nhc.gov.cn/xcs/yqtb/202002/553ff43ca29d4fe88f3837d49d6b6ef1.shtml (accessed 14 Feb 2020.

7. World Health Organization. Novel Coronavirus (2019-nCoV) Situation Report - 24, 2020. 2020. https://www.who.int/docs/default-source/coronaviruse/situation-reports/20200213-sitrep-24-covid-19.pdf (accessed 14 Feb 2020.

8. World Health Organization. Infection prevention and control during health care when novel coronavirus (nCoV) infection is suspected. 2020. https://www.who.int/publications-detail/infection-prevention-and-control-during-health-care-when-novel-coronavirus-(ncov)-infection-is-suspected-20200125 (accessed 7 Feb 2020.

9. Ministry of Industry and Information Technology of People ‘s Republic of China. Facemasks shortage? Just now, the Ministry of Industry and Information Technology of People ‘s Republic of China responded (in Chinese). 2020. http://www.miit.gov.cn/n973401/n7647394/n7647409/c7656383/content.html (accessed 7 Feb 2020.

10. Ming W, Huang J, Zhang C. Breaking down of the healthcare system: Mathematical modelling for controlling the novel coronavirus (2019-nCoV) outbreak in Wuhan, China. bioRxiv 2020.

11. Jefferson T, Del Mar C, Dooley L, et al. Physical interventions to interrupt or reduce the spread of respiratory viruses: systematic review. BMJ 2009; 339: b3675.

12. World Health Organization (WHO). WHO Director-General’s opening remarks at the media briefing on 2019 novel coronavirus – 7 February 2020. 2020. https://www.who.int/dg/speeches/detail/who-director-general-s-opening-remarks-at-the-media-briefing-on-2019-novel-coronavirus7-february-2020 (accessed 14 Feb 2020.

13. Ministry of Industry and Information Technology of People ‘s Republic of China. Press conference on the situation of medical protective equipment during the outbreak of 2019-nCoV pneumonia (in Chinese). 2020. http://www.miit.gov.cn/n1146290/n1146402/c7660995/content.html (accessed 7 Feb 2020.

14. The General Administration of Customs of the People’s Republic of China. China has imported 730 million facemasks since the New Year’s eve (in Chinese). 2020. http://www.gov.cn/xinwen/2020-02/12/content_5477838.htm (accessed 14 Feb 2020.

15. Ministry of Industry and Information Technology of People ‘s Republic of China. A large numbers of clothing enterprises changed to produce facemasks and protective clothes. 2020. http://www.miit.gov.cn/n973401/n7647394/n7647409/c7675791/content.html (accessed 14 Feb 2020.

16. Kucharski A, Russell T, Diamond C, et al. Analysis and projections of transmission dynamics of nCoV in Wuhan. 2020. https://cmmid.github.io/ncov/wuhan_early_dynamics/index.html (accessed 14 Feb 2020.

17. Liu Q, Li D, Liu Z, et al. EPIDEMIC TRENDS ANALYSIS AND RISK ESTIMATION OF 2019-NCOV OUTBREAK. bioRxiv 2020.

18. National Health Commission of the People ‘s Republic of China. Chinese Health Statistics Yearbook (2019 version) (in Chinese). China; 2019.

19. National Bureau of Statistics of China. Main cities annual statistical data (in Chinese). 2019. http://data.stats.gov.cn/easyquery.htm?cn=E0105&zb=A02&reg=110000&sj=2018 (accessed 14 Feb 2020.

20. Wuhan Bureau of Statistics. Wuhan statistical bulletin on national economic and social development in 2018 (in Chinese). 2019. http://www.tjcn.org/tjgb/17hb/35881.html (accessed 14 Feb 2020.

21. National Health Commission of the People ‘s Republic of China. The protocol for the novel coronavirus pneumonia (the second version) (in Chinese). 2020. http://www.nhc.gov.cn/jkj/s3577/202001/c67cfe29ecf1470e8c7fc47d3b751e88.shtml (accessed 7 Feb 2020.

22. Ministry of Education of the People’s Republic of China. The announcement on the postponement of the spring semester of 2020. 2020. http://www.moe.gov.cn/jyb_xwfb/gzdt_gzdt/s5987/202001/t20200127_416672.html (accessed 17 Feb 2020.

23. World Heath Organization. WHO, China leaders discuss next steps in battle against coronavirus outbreak. 2020. https://www.who.int/news-room/detail/28-01-2020-who-china-leaders-discuss-next-steps-in-battle-against-coronavirus-outbreak (accessed 7 Feb 2020.

24. The General Administration of Customs of the People’s Republic of China. Proper reservation and disinfection methods could prolong the using time and times of facemasks (in Chinese). 2020. http://www.gov.cn/xinwen/2020-02/06/content_5475327.htm (accessed 15 Feb 2020.

25. Song W, Pan B, Kan H, Xu Y, Yi Z. Evaluation of heat inactivation of virus contamination on medical mask. Journal of Microbes and Infections 2020.

26. Ministry of Industry and Information Technology of People ‘s Republic of China. Chinese medical N95 mask daily productivity is only 600,000! Forward the appeal, please leave it to the medical staff (in Chinese). 2020. http://www.miit.gov.cn/n973401/n7647394/n7647409/c7663732/content.html (accessed 7 Feb 2020.

27. Reynolds DL, Garay JR, Deamond SL, Moran MK, Gold W, Styra R. Understanding, compliance and psychological impact of the SARS quarantine experience. Epidemiol Infect 2008; 136(7): 997–1007.

28. CNBC. Panic buying of face masks is unwarranted and could pose risks for health workers, experts say. 2020. https://www.cnbc.com/2020/01/31/china-coronavirus-shortage-of-face-masks-could-pose-risks-for-healthcare-workers.html (accessed 7 Feb 2020.

29. National Health Commission of the People ‘s Republic of China. The announcement of publishing the guidelines of protection of population at different risk levels for novel coronavirus and facemasks usage to prevent the novel coronavirus infection (in Chinese). 2020. http://www.nhc.gov.cn/jkj/s7916/202001/a3a261dabfcf4c3fa365d4eb07ddab34.shtml (accessed 7 Feb 2020.

30. Chinanews.com. Feb 21 maybe peak day for coronavirus outbreak: Yale expert. 2020. http://www.ecns.cn/news/2020-02-04/detail-ifztewca0596265.shtml (accessed 7 Feb 2020.

31. Wu JT, Leung K, Leung GM. Nowcasting and forecasting the potential domestic and international spread of the 2019-nCoV outbreak originating in Wuhan, China: a modelling study. Lancet 2020.

